# Comparative 48-Week Viral Load Suppression across Antiretroviral Initiation Regimens: Dolutegravir versus Non-Dolutegravir among People Living with HIV in Tanzania

**DOI:** 10.64898/2026.03.19.26348839

**Authors:** Gwandumi F. Kayange, Raphael Z. Sangeda, Prosper Njau

## Abstract

**Background:** Routine viral load monitoring is central to assessing treatment effectiveness in HIV care, and dolutegravir (DTG)-based regimens are now preferred in many treatment programmes. However, national routine data analyses comparing 48-week viral load suppression across antiretroviral therapy initiation regimens in Tanzania remain limited.

**Methods:** We conducted a retrospective cohort analysis using routinely collected HIV programme data from Tanzania’s National AIDS, STIs and Hepatitis Control Programme database. After de-duplication and data processing, the working analysis warehouse contained 49,547 patients and 1,008,137 visits. The primary analysis included 6,991 patients with a valid viral load measured 48 weeks after initiation of antiretroviral therapy. Viral suppression was defined as a viral load <1,000 copies/mL. We compared suppression between DTG-based and non-DTG-based initiation groups and across individual initiation regimens. Treatment change episodes and early DTG switching patterns were summarized as secondary analyses.

**Results:** Of the 6,991 included patients, 6,113 (87.4%) achieved viral load suppression at 48 weeks. Suppression was higher among DTG initiators than non-DTG initiators (917/1,000, 91.7% vs. 5,196/5,991, 86.7%). TDF+3TC+EFV was the most common non-DTG initiation regimen, whereas TDF+3TC+DTG was the most common regimen among DTG initiators.

**Conclusions:** Viral suppression at 48 weeks was high overall but was higher among patients initiated on DTG-based regimens than among those initiated on non-DTG regimens. By anchoring outcomes to a fixed post-initiation time point, this study complements existing Tanzanian evidence on viral load testing uptake and geographic variation. It provides regimen-specific insights into the effectiveness of early treatment under routine programme conditions.

## Introduction

HIV remains a major global public health challenge despite substantial progress in diagnosis, treatment, and survival. Expanded access to antiretroviral therapy has transformed HIV into a manageable chronic condition for many people; however, sustained viral suppression remains central to both individual clinical benefits and prevention of onward transmission [1–3].

Routine viral load monitoring is now a core component of HIV programme performance in sub-Saharan Africa. Although monitoring capacity has expanded considerably, important gaps remain in terms of access, timeliness, and equity. Viral load testing provides both patient-level evidence of treatment effectiveness and programme-level insight into areas where treatment delivery may be underperforming [4,5]. At the same time, the interpretation of programme performance depends not only on whether testing occurs but also on how virologic outcomes relate to treatment choices.

Dolutegravir (DTG) has become central to contemporary HIV treatment policies. The World Health Organization recommends DTG as the preferred first- and second-line HIV treatment option because of its effectiveness, tolerability, and high genetic barrier to resistance [6,7]. Accumulating evidence from observational cohorts and clinical trials has demonstrated strong virologic outcomes with DTG-based therapy in both treatment-naïve and treatment-experienced populations [8–12].

In Tanzania, DTG-based regimens have been incorporated into routine HIV care, and recent national analyses have described viral load testing uptake, suppression among those tested, and geographic heterogeneity in programme performance [13,14]. However, these analyses are largely cross-sectional or programme-level and do not directly evaluate virologic outcomes at a fixed time point after treatment initiation.

This distinction is important. A cohort-based 48-week analysis anchors virologic outcomes to a clinically meaningful post-initiation milestone, thereby reducing temporal variability introduced by programme scale-up and evolving treatment policies. In contrast to cross-sectional assessments of viral load uptake or suppression, this approach allows a more direct comparison of the effectiveness of early treatment across initiation regimens, particularly during transitions from non-DTG to DTG-based therapy.

Therefore, we aimed to compare 48-week viral load suppression across antiretroviral therapy initiation regimens, focusing on DTG versus non-DTG, among people living with HIV in Tanzania. As a secondary objective, we described treatment change episodes and early switching patterns during the first 48 weeks of follow-up to provide context on treatment dynamics during the transition to DTG-based care.

## Methods

### Study design and data source

We conducted a retrospective cohort analysis using routinely collected data from the Tanzania National AIDS Sexually Transmitted Infections and Hepatitis Control Programme (NASHCoP) Care and Treatment Clinic (CTC) database. The database contains longitudinal patient-level records from HIV care and treatment facilities across mainland Tanzania.

### Cohort construction and sampling

The national database was first de-duplicated at the patient level to ensure unique identifiers across facilities. From the de-duplicated national dataset, a simple random sample of 50,000 unique patient identifiers was generated using R.

The sampled patient IDs were then used to extract linked records across multiple datasets, including baseline demographic data, antiretroviral therapy (ART) regimens, clinic visits, laboratory results, and anthropometric measurements. These datasets were merged to form a longitudinal analytical dataset containing 49,872 individuals and over one million clinical visits.

### Study population

For the primary analysis, we included individuals with a documented viral load measurement approximately 48 weeks (± allowable programmatic window based on routine testing schedules). After applying this criterion, 6,991 individuals were included in the final analytic cohort.

### Exposure definition

The primary exposure was an ART regimen at initiation. The regimens were classified into DTG-based regimens (initiation regimens containing DTG) and non-DTG regimens (NNRTI-based and other non-DTG regimens). Patient classifications were based on standardized regimen coding derived from national treatment records.

### Regimen coding and harmonization

Regimen components were coded using standard antiretroviral abbreviations: TDF (tenofovir disoproxil fumarate), FTC (emtricitabine), 3TC (lamivudine), EFV (efavirenz), DTG (dolutegravir), ABC (abacavir), NVP (nevirapine), ATV/r (atazanavir/ritonavir), and LPV/r (lopinavir/ritonavir). Regimen codes from the national database were harmonized into standardized regimen names to enable consistent classification and comparison across treatment groups and calendar years.

### Outcome definition

The primary outcome was viral load suppression at 48 weeks after ART initiation, defined as a viral load <1,000 copies/mL. A binary variable was constructed as 1 for suppressed (<1,000 copies/mL) and 0 for not suppressed (≥1,000 copies/mL).

### Covariates

The following covariates were included based on clinical relevance and data availability: age at ART initiation (continuous). gender(male, female). average adherence (percentage) and calendar year of ART initiation (2017–2021).

### Reconstruction of Longitudinal Clinical Trajectories

Longitudinal treatment histories were reconstructed from electronic medical records using a customized data engineering pipeline. A Treatment Change Episode (TCE) was defined as the transition from a non-DTG regimen to a DTG-based backbone. To evaluate medication adherence, we calculated pharmacy refill [15,16] adherence as a continuous variable derived from scheduled appointment intervals and actual visit dates. Population-level transitions were visualized using sorted Swimmer Plots (n=40). At the same time, individual-level clinical outcomes were assessed using 4-panel integrated trajectories that synchronize regimen changes with longitudinal adherence, viral load (log scale), and CD4 count.

### Statistical analysis

Descriptive analyses summarized cohort characteristics overall and by regimen group. Continuous variables are reported as means with standard deviations, and categorical variables as counts and percentages.

Crude comparisons of viral load suppression between the DTG and non-DTG groups were performed.

Multivariate logistic regression was used to assess the association between regimen type and 48-week viral load suppression, adjusting for age, gender, adherence, and the calendar year of ART initiation. The calendar year was modeled as a categorical variable to account for programmatic changes over time. A sensitivity analysis was conducted, modeling the year as a continuous variable.

### Additional analyses

To assess potential temporal confounding related to the scale-up of DTG-based therapy, we examined the distribution of ART initiation by calendar year, trends in DTG uptake over time, and changes in viral load suppression across calendar years.

Regimen-level analyses were additionally conducted using harmonized regimen classifications derived from national coding systems and are presented in the Supplementary Materials. These analyses were intended to distinguish regimen effects from programme-level changes occurring during the transition to DTG-based care.

## Results

### Study population

A total of 6,991 individuals with a documented viral load measurement approximately 48 weeks after ART initiation were included in the analysis. The mean age at ART initiation was 37.6 years (SD 13.8), and the mean adherence was 86.9% (SD 12.2). Patients had an average of 26.2 clinic visits (SD 11.7) during follow-up.

### Baseline characteristics

Baseline characteristics differed between patients initiating DTG-based and non-DTG regimens (Table 1). Patients initiating DTG-based regimens were slightly younger (36.7 vs 37.8 years; p=0.012) and had lower mean adherence (84.6% vs 87.3%; p<0.001). They also had fewer clinic visits (15.7 vs 28.0; p<0.001) and shorter follow-up duration (627.0 vs 1365.5 days; p<0.001).

DTG initiators had fewer therapy changes (mean 0.3 vs 1.4; p<0.001) and fewer treatment change episodes (1.5 vs 3.0; p<0.001). Sex distribution was similar between groups (p=0.212).

ART initiation year differed substantially between regimen groups, reflecting programme transition. Most non-DTG initiators started treatment in 2017–2018, whereas DTG initiators were concentrated in later years, particularly 2020–2021. These differences reflect the phased introduction and scale-up of DTG-based regimens within the national programme.

**Table 1.** Baseline characteristics of the study population according to ART initiation regimen (DTG vs. non-DTG).

| **Variable** | **Overall** | **Non-DTG** | **DTG** | **P-value** |
| --- | --- | --- | --- | --- |
| Age at ART initiation (years) | 37.6 (13.8) | 37.8 (14.1) | 36.7 (12.0) | 0.012 |
| Adherence (%) | 86.9 (12.2) | 87.3 (11.8) | 84.6 (14.4) | <0.001 |
| Number of clinic visits | 26.2 (11.7) | 28.0 (11.3) | 15.7 (7.6) | <0.001 |
| Follow-up duration (days) | 1259.8 (498.4) | 1365.5 (440.2) | 627.0 (329.3) | <0.001 |
| Therapy change count | 1.2 (0.9) | 1.4 (0.8) | 0.3 (0.6) | <0.001 |
| Treatment change episode count | 2.8 (3.2) | 3.0 (3.4) | 1.5 (1.4) | <0.001 |
| Female | 4509 (64.5%) | 3882 (64.8%) | 627 (62.7%) | 0.212 |
| Male | 2482 (35.5%) | 2109 (35.2%) | 373 (37.3%) |  |
| ART initiation year: 2017 | 3947 (56.5%) | 3890 (64.9%) | 57 (5.7%) | <0.001 |
| ART initiation year: 2018 | 1111 (15.9%) | 1080 (18.0%) | 31 (3.1%) |  |
| ART initiation year: 2019 | 1000 (14.3%) | 762 (12.7%) | 238 (23.8%) |  |
| ART initiation year: 2020 | 858 (12.3%) | 258 (4.3%) | 600 (60.0%) |  |
| ART initiation year: 2021 | 75 (1.1%) | 1 (0.0%) | 74 (7.4%) |  |

Note: Values are presented as mean (SD) for continuous variables and n (%) for categorical variables. P-values compare DTG and non-DTG groups.

### Temporal distribution of DTG initiation

ART initiation patterns changed markedly over calendar time. In 2017, DTG-based regimens accounted for only 57 of 3,947 initiations (1.4%), increasing to 31 of 1,111 (2.8%) in 2018, 238 of 1,000 (23.8%) in 2019, and rising sharply to 600 of 858 (70.0%) in 2020 and 74 of 75 (98.7%) in 2021. These trends indicate a rapid transition from non-DTG to DTG-based therapy during the study period (Figure 1).

**Figure 1.**
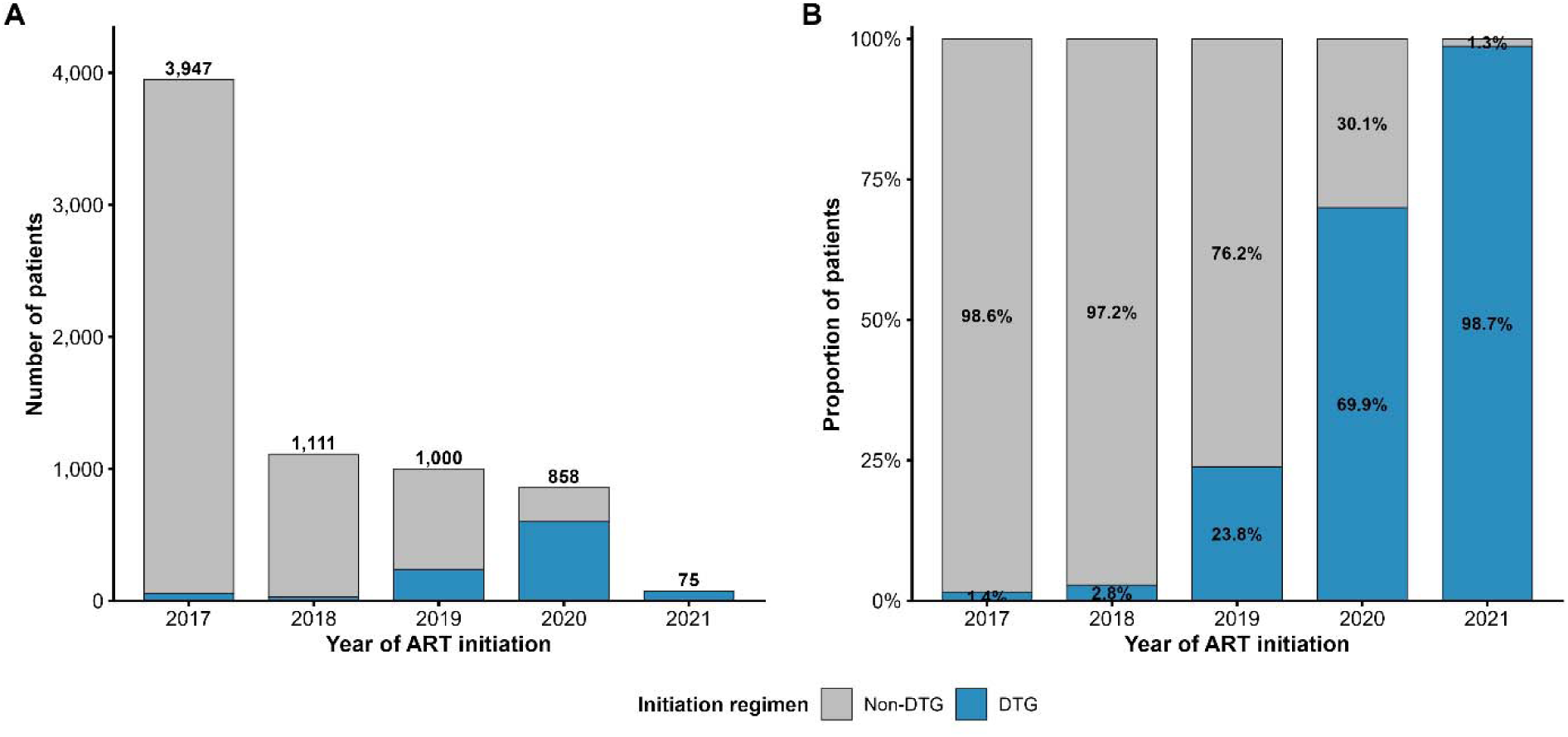
Distribution of ART initiation regimens by calendar year. **Panel A** shows the number of patients initiating DTG-based and non-DTG regimens by year. **Panel B** shows the proportion of initiations accounted for by each regimen group within each calendar year.

### Crude viral load suppression

At 48 weeks after ART initiation, overall viral load suppression was 6,113 of 6,991 (87.4%). Suppression was higher among patients initiated on DTG-based regimens compared with those initiated on non-DTG regimens (917 of 1,000, 91.7% vs 5,196 of 5,991, 86.7%) (Figure 2).

**Figure 2.**
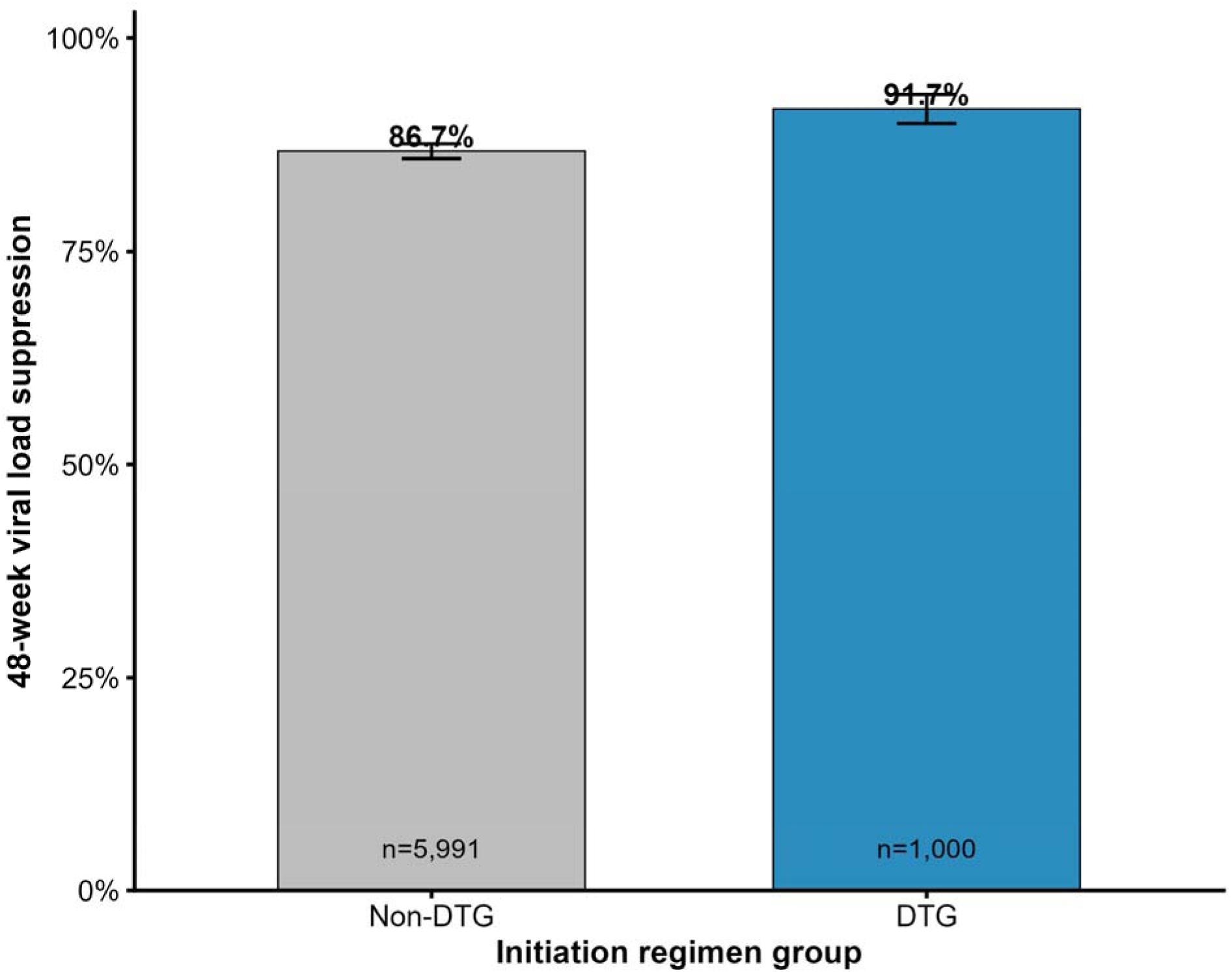
Crude 48-week viral load suppression by ART initiation regimen. Bars represent the proportion of patients achieving viral load suppression (<1,000 copies/mL) at 48 weeks among DTG-based and non-DTG initiation groups.

### Regimen-level viral load suppression

Regimen-specific 48-week viral load suppression estimates are presented in Supplementary Tables S1 and S2. The most common non-DTG regimen, TDF+3TC+EFV, demonstrated high suppression (89.7%), while TDF+3TC+DTG achieved 91.8% suppression overall. Across calendar years, suppression improved within both regimen groups, with DTG-based regimens showing consistently high suppression in later years. Small sample sizes limit the interpretation of less frequent regimens.

### Multivariate analysis

In the adjusted logistic regression model, DTG-based initiation was not associated with higher odds of 48-week viral load suppression compared with non-DTG initiation (adjusted OR 0.98, 95% CI 0.71–1.37; p=0.924) (Table 2).

**Table 2.** Multivariate logistic regression analysis of factors associated with 48-week viral load suppression.

| Variable | Adjusted OR | 95% CI | P-value |
| --- | --- | --- | --- |
| DTG vs Non-DTG | 0.98 | 0.71–1.37 | 0.924 |
| Age at ART initiation (per year) | 1.02 | 1.02–1.03 | <0.001 |
| Male vs Female | 0.74 | 0.64–0.87 | <0.001 |
| Adherence (per % increase) | 1.02 | 1.02–1.03 | <0.001 |
| Year 2018 vs 2017 | 1.93 | 1.56–2.41 | <0.001 |
| Year 2019 vs 2017 | 2.81 | 2.17–3.70 | <0.001 |
| Year 2020 vs 2017 | 2.72 | 1.93–3.88 | <0.001 |
| Year 2021 vs 2017 | 2.22 | 1.00–5.67 | 0.068 |

Note: Adjusted for age at ART initiation, sex, adherence, and calendar year of ART initiation. Odds ratios (ORs) with 95% confidence intervals (CI) are shown.

Older age at ART initiation (adjusted OR 1.02 per year, 95% CI 1.02–1.03; p<0.001) and higher adherence (adjusted OR 1.02 per percentage increase, 95% CI 1.02–1.03; p<0.001) were associated with increased odds of suppression, whereas male sex was associated with lower odds of suppression (adjusted OR 0.74, 95% CI 0.64–0.87; p<0.001).

Calendar year of ART initiation was strongly associated with improved suppression, with higher odds observed in later years compared with 2017, particularly in 2019 (adjusted OR 2.81, 95% CI 2.17–3.70; p<0.001) and 2020 (adjusted OR 2.72, 95% CI 1.93–3.88; p<0.001).

### Temporal trends in viral load suppression

Crude viral load suppression increased over calendar time in both regimen groups. Suppression was lower in earlier years and improved substantially by 2019–2020, with similar levels observed between DTG and non-DTG groups in later years (Figure 3).

**Figure 3.**
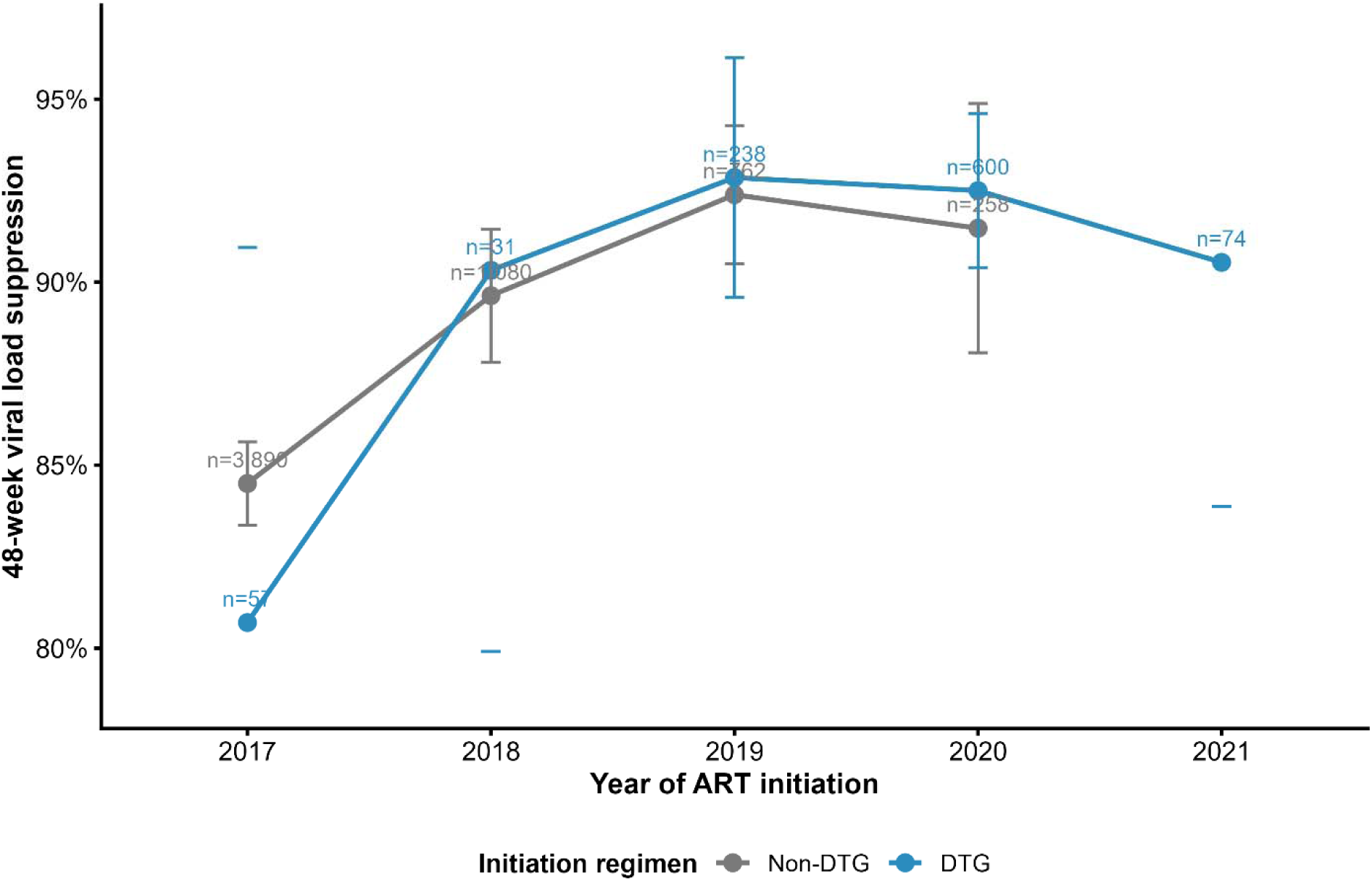
Crude 48-week viral load suppression by calendar year of ART initiation, stratified by regimen group. Points represent the proportion of patients achieving viral load suppression (<1,000 copies/mL) at 48 weeks within each calendar year and regimen group. Error bars indicate 95% confidence intervals. Subgroups with very small sample sizes (n < 10) were excluded.

### Programmatic Optimization Pathways and Virologic Rescue

Among patients undergoing a Treatment Change Episode, the predominant optimization pathway was the transition from TDF+3TC+EFV to TDF+3TC+DTG (69.4%), followed by AZT+3TC+NVP to TDF+3TC+DTG (10.4%). Swimmer plot analysis (Figure 4) demonstrated that programmatic optimization was rapidly scalable, with most transitions occurring within 6–9 months of the national rollout.

**Figure 4:**
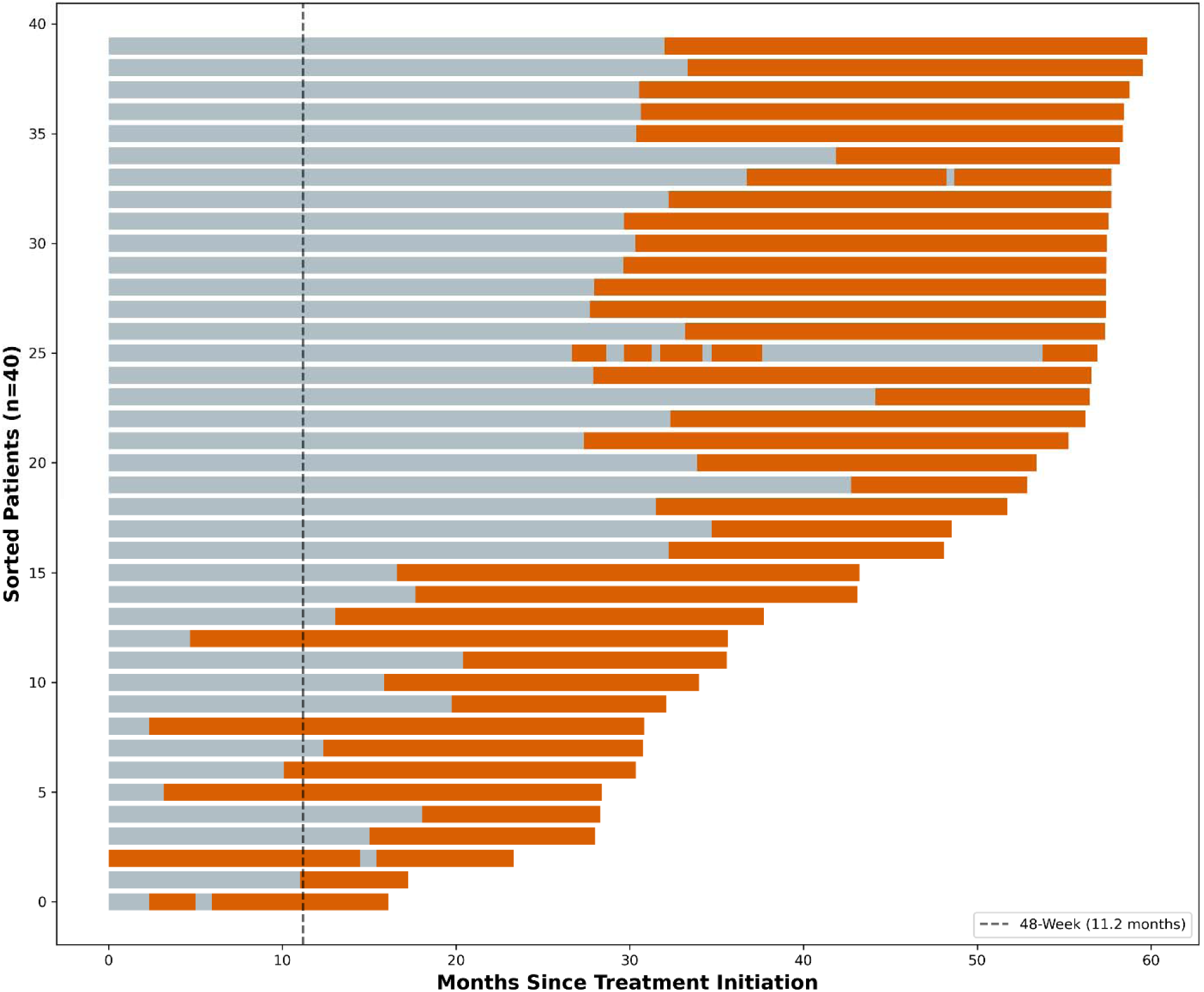
Population-Level Treatment Change Episodes (TCE) Swimmer Plot of Longitudinal Treatment Histories Among Early DTG-Switchers (n=40). Each horizontal bar represents the treatment journey of an individual patient, sorted by the total follow-up duration. Grey segments indicate periods on non-DTG-based regimens (primarily NNRTI-based), while orange segments represent the transition to DTG-based therapy. The vertical dashed line indicates the 48-week (11.2 months) landmark. The plot illustrates the rapid and systematic programmatic scale-up of the DTG transition within the national cohort, with the majority of “early switchers” successfully optimized within the first 6–9 months of follow-up.

Case-level longitudinal analysis (Figure 5) revealed that virologic failure (>1,000 copies/mL) on initial NNRTI-based regimens was frequently preceded by significant dips in pharmacy-refill adherence. Notably, the transition to DTG-based therapy resulted in rapid and sustained virologic suppression. This “virologic rescue” occurred even in patients with a history of inconsistent adherence, highlighting the high genetic barrier to resistance and potency of the DTG backbone in this real-world Tanzanian cohort.

**Figure 5:**
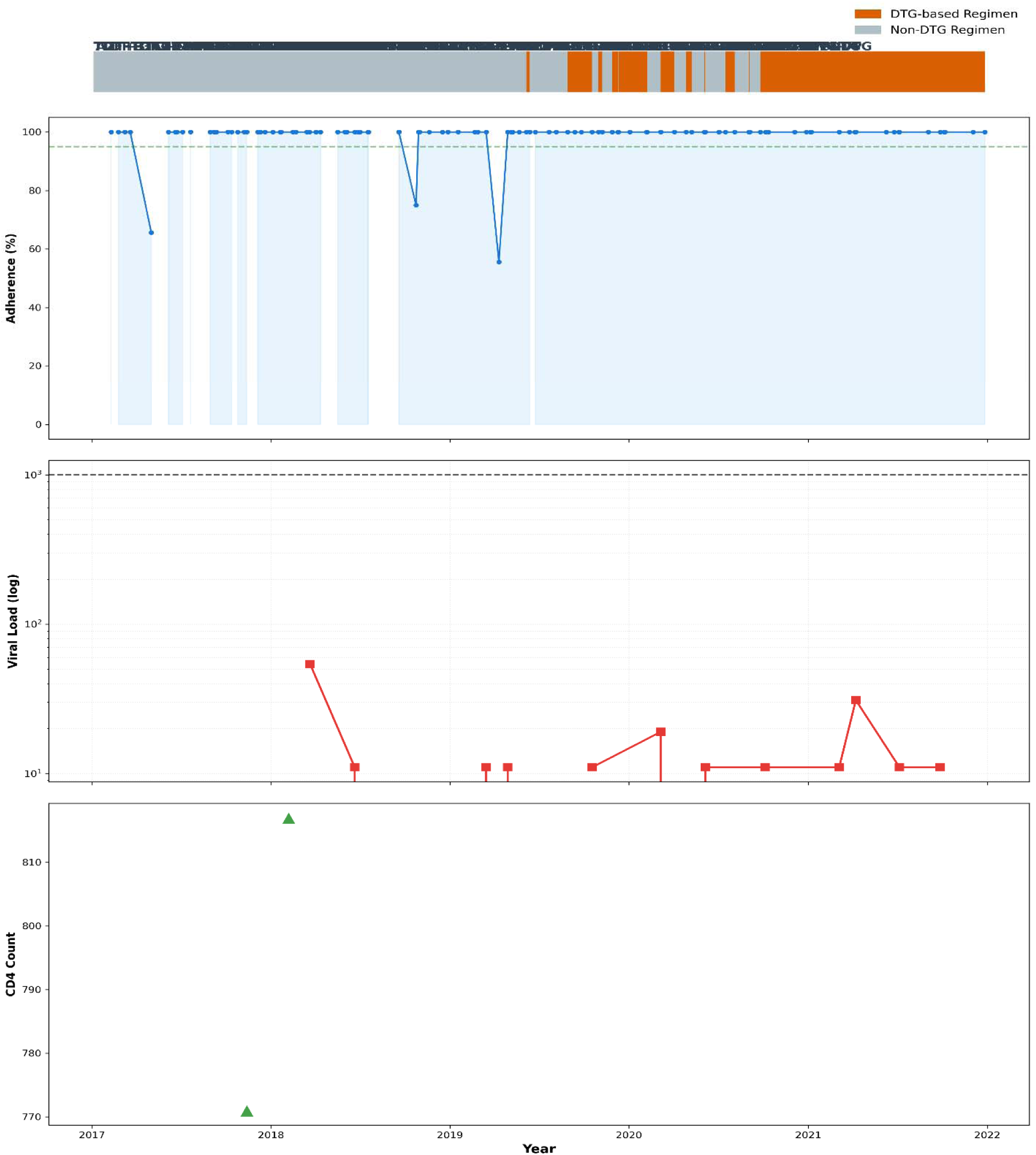
Integrated Clinical Trajectory. **Longitudinal Correlation of Medication Adherence, Regimen Optimization, and Virologic Outcomes.** This representative case study displays four synchronized clinical dimensions over time: **(A) Regimen Timeline:** showing the transition from a non-DTG regimen (grey) to a DTG-based regimen (orange); **(B) Pharmacy Adherence:** percentage of adherence based on pharmacy refill intervals, showing a significant decline preceding virologic failure; **(C) Viral Load Trajectory:** longitudinal viral load (log scale) showing a failure spike (>1,000 copies/mL) during sub-optimal adherence on the initial regimen, followed by rapid and sustained suppression post-DTG transition; and **(D) Immunologic Recovery:** CD4 count recovery trend (cells/mm³) following the Treatment Change Episode (TCE).

## Discussion

In this national routine-data analysis of people living with HIV in Tanzania with documented viral load results at 48 weeks after the initiation of antiretroviral therapy, overall suppression was high, with 87.4% of patients achieving a viral load <1,000 copies/mL. Suppression was higher among those initiated on DTG-based regimens than among those initiated on non-DTG-based regimens (91.7% vs. 86.7%, respectively). These findings indicate that DTG-based initiation was associated with higher crude short-term virological outcomes at a clinically meaningful follow-up milestone. However, this apparent advantage was attenuated after adjustment for adherence and calendar year of ART initiation, suggesting that temporal programme scale-up and adherence differences contributed to the observed crude association.

This pattern is consistent with the broader evidence base supporting DTG as a preferred component of contemporary HIV treatment. The World Health Organization recommends DTG as the preferred first- and second-line option due to its potency, tolerability, and high barrier to resistance [6,7]. Observational studies and randomized trials have demonstrated strong virologic performance of DTG-based regimens in both treatment-naïve and treatment-experienced populations [8–12,17]. Therefore, the higher suppression observed in this analysis is consistent with the expected programmatic advantages under routine care conditions. These data support the biological and clinical advantages of DTG-based therapy but do not exclude the influence of programme-level factors on observed outcomes in routine care settings.

Randomized evidence from the D2EFT trial further demonstrated that DTG-containing regimens achieve high rates of virologic suppression even in second-line settings following NNRTI failure, supporting the durability and robustness of DTG-based therapy across treatment lines [17].

Our findings provide robust real-world evidence of the effectiveness of the Tanzanian national DTG transition strategy. The heavy reliance on the TDF+3TC+EFV to TDF+3TC+DTG pathway (69.4%) aligns with World Health Organization (WHO) recommendations for optimizing first-line therapy [18,19]. The observed rapid virologic suppression post-TCE, despite prior adherence challenges, suggests that the transition served as a critical corrective intervention for patients struggling with NNRTI-based regimens. These data underscore the utility of the DTG-based TCE as a primary tool for achieving and sustaining the UNAIDS 95-95-95 targets in resource-limited settings [20,21].

At the same time, the findings do not suggest poor performance of non-DTG regimens. The non-DTG group achieved a high 48-week suppression rate of 86.7%, and the dominant regimen, TDF+3TC+EFV, achieved 89.7% suppression. This is important in the Tanzanian context, where many patients initiated treatment during a period when efavirenz-based regimens were widely used and where the transition to DTG occurred progressively. This reinforces that high levels of virologic suppression were achievable under efavirenz-based regimens within routine programme conditions prior to full DTG scale-up. These findings align with national evidence demonstrating strong viral suppression in routine programme populations [13,14,22,23], while extending prior work by applying a fixed 48-week post-initiation framework rather than cross-sectional assessments of suppression.

The distribution of regimens in this cohort reflects the evolution of national treatment policy. Most patients initiated on non-DTG regimens, particularly TDF+3TC+EFV, whereas DTG initiators predominantly received TDF+3TC+DTG. This pattern captures the temporal transition in prescribing practices during the national scale-up of DTG. The loss of statistical significance after adjustment indicates that calendar time and programme maturation, rather than regimen alone, largely explained the observed crude differences. Therefore, the comparison between regimen groups should be interpreted within this historical context, in which differences partly reflect both regimen efficacy and changing programme implementation over time.

In addition, virologic outcomes in routine programme settings are strongly influenced by patient-level factors, particularly adherence and baseline clinical status [4,24]. In this analysis, adherence was independently associated with viral load suppression, reinforcing its central role in treatment success. Variability in baseline disease severity, including advanced HIV disease, may also contribute to differences in observed outcomes across patient groups. The relative effectiveness of dolutegravir-based regimens may therefore not be uniform across all patients, but instead vary depending on clinical context. These factors are not fully captured in routine programme datasets but are important in interpreting regimen-level effectiveness beyond the choice of antiretroviral agents alone [25].

Treatment change episodes were common, supporting their relevance as a secondary analytical dimension. In routine HIV care, regimen changes may occur due to optimization, tolerability, simplification, evolving guidelines, or virologic considerations [9,26]. In this dataset, such changes are likely also reflective of a programmatic transition toward DTG-based therapy. Findings from related Tanzanian analyses have shown that treatment continuity and outcomes are closely linked to adherence patterns and programmatic dynamics [15,22,27]. Once the date-based switching analysis is fully incorporated, it will provide additional insights into early transitions from non-DTG to DTG regimens during the first 48 weeks and clarify which initial regimens most frequently preceded switching. These findings are consistent with the observed differences in treatment change episode counts between regimen groups in this cohort.

Together, these findings highlight that early virologic outcomes in routine HIV care reflect not only the choice of antiretroviral regimen, but also the timing of programme implementation and patient-level factors, underscoring the need for context-aware interpretation of treatment effectiveness.

This study has several strengths. It uses national routine programme data, includes a large analytical warehouse, and focuses on a clinically interpretable 48-week endpoint that aligns with treatment monitoring practices. Importantly, it addresses a distinct analytical question compared with prior Tanzanian work by anchoring outcomes to a fixed post-initiation time point rather than examining aggregate suppression or testing-uptake patterns [14]. This design reduces temporal misclassification and provides a clearer comparison of early treatment effectiveness across initiation regimens.

However, several limitations should be considered. First, only 6,991 of 49,547 patients in the analytic dataset had a valid 48-week viral load, raising the possibility of selection bias. Patients with documented 48-week viral load measurements may differ systematically from those without such measurements, particularly with respect to retention, adherence, or access to testing. In addition, residual confounding by unmeasured factors, including baseline clinical status and facility-level differences, cannot be excluded. Second, some regimen categories had small sample sizes, limiting the stability and interpretability of the individual-regimen estimates. Finally, the analysis is observational and descriptive; therefore, differences between regimen groups should not be interpreted as causal without adjustment for potential confounders.

In conclusion, this study demonstrates high overall viral load suppression at 48 weeks among people living with HIV in Tanzania, with higher crude suppression observed among patients initiated on DTG-based regimens compared with non-DTG regimens. However, this difference was not sustained after adjustment for adherence and calendar year, highlighting the importance of programme-level factors in interpreting regimen effects. These findings support the continued programmatic value of DTG-based therapy and highlight the substantial effectiveness of non-DTG regimens, particularly TDF+3TC+EFV, during the transition period. Completion of the switching analysis will further strengthen the interpretation by providing insights into treatment dynamics during DTG scale-up in Tanzania.

## Data Availability

The data used in this study were derived from routine national HIV programme records and are not publicly available. Access is subject to approval from the National AIDS, STIs and Hepatitis Control Programme and relevant Tanzanian authorities.

## Declarations

### Ethics approval and consent to participate

This study used routinely collected HIV programme data from the National AIDS, STIs and Hepatitis Control Programme database in Tanzania. Ethical and administrative approvals for the use of programme data were obtained from the relevant institutional and national authorities. Patient confidentiality was maintained throughout, and no directly identifying information was included in the analytic dataset or manuscript with reference number DA.228/298/02L/502, dated February 25, 2025.

### Consent for publication

Not applicable. This manuscript does not include identifiable individual-level data.

## Authors’ contributions

GFK contributed to data interpretation, preliminary analysis, manuscript drafting, and preparation of tables and figures. RZS conceived the study, supervised the work, guided the analytical framework, interpreted the findings, and critically revised the manuscript for intellectual content. PN contributed to programme-level interpretation, contextualization of the data, and critical review of the manuscript.

## Acknowledgements

We acknowledge the National AIDS, STIs and Hepatitis Control Programme for providing access to routine programme data. We also recognize the contributions of healthcare workers involved in HIV care and data recording across Tanzania.

