## Supplementary Materials for "Comparative 48-Week Viral Load Suppression across Antiretroviral Initiation Regimens: Dolutegravir versus Non-Dolutegravir among People Living with HIV in Tanzania"

Detailed regimen-level outcomes for 48-week viral load suppression using harmonized regimen classifications are presented in Supplementary Tables S1 and S2. Regimen codes from the national database were consolidated into standardized regimen names to enable meaningful comparison across treatment groups.

Most patients initiated TDF-based regimens, while DTG-based regimens were concentrated in later calendar years, reflecting the temporal transition in national treatment policy.

**Supplementary Table S1:** Regimen-level 48-week viral load suppression among patients initiating antiretroviral therapy, presented using harmonized regimen names.

| Regimen | n | Suppressed (n) | Suppression (%) |
|---|---|---|---|
| TDF+3TC+EFV | 4,554 | 4,087 | 89.7 |
| TDF+3TC+DTG | 985 | 904 | 91.8 |
| AZT+3TC+NVP | 720 | 536 | 74.4 |
| AZT+3TC+EFV | 292 | 228 | 78.1 |
| TDF+FTC+EFV | 218 | 192 | 88.1 |
| ABC+3TC+EFV | 69 | 53 | 76.8 |
| ABC+3TC+LPV/r | 69 | 48 | 69.6 |
| TDF+FTC+ATV/r | 23 | 16 | 69.6 |
| TDF+FTC+LPV/r | 20 | 19 | 95.0 |
| ABC+3TC+DTG | 14 | 12 | 85.7 |
| AZT+3TC+LPV/r | 8 | 5 | 62.5 |
| ABC+3TC+ATV/r | 7 | 4 | 57.1 |
| AZT+3TC+ATV/r | 5 | 3 | 60.0 |
| Other 2nd line | 2 | 1 | 50.0 |
| TDF+3TC+NVP | 2 | 2 | 100 |
| TDF+FTC+NVP | 2 | 2 | 100 |
| AZT+3TC+DTG | 1 | 1 | 100 |

**Supplementary Table S2:** Regimen-specific 48-week viral load suppression stratified by calendar year of ART initiation (wide format).

| Regimen | 2017 (%) | 2018 (%) | 2019 (%) | 2020 (%) | 2021 (%) | 2017 n | 2018 n | 2019 n | 2020 n | 2021 n |
|---|---|---|---|---|---|---|---|---|---|---|
| TDF+3TC+EFV | 88.1 | 90.7 | 93.3 | 93.4 | 100 | 2634 | 974 | 716 | 229 | 1 |
| TDF+3TC+DTG | 81.5 | 90.3 | 93.2 | 92.4 | 90.5 | 54 | 31 | 234 | 592 | 74 |
| AZT+3TC+NVP | 74.0 | 73.7 | 78.3 | 100 | NA | 631 | 57 | 23 | 9 | NA |
| AZT+3TC+EFV | 77.8 | 100 | NA | NA | NA | 288 | 4 | NA | NA | NA |
| TDF+FTC+EFV | 88.3 | 86.7 | 100 | 50 | NA | 197 | 15 | 4 | 2 | NA |
| ABC+3TC+EFV | 73.7 | 85.7 | 85.7 | 33.3 | NA | 38 | 21 | 7 | 3 | NA |
| ABC+3TC+LPV/r | 65.7 | 87.5 | 63.6 | 73.3 | NA | 35 | 8 | 11 | 15 | NA |
| TDF+FTC+ATV/r | 68.2 | 100 | NA | NA | NA | 22 | 1 | NA | NA | NA |
| TDF+FTC+LPV/r | 94.7 | NA | 100 | NA | NA | 19 | NA | 1 | NA | NA |
| ABC+3TC+DTG | 50.0 | NA | 75.0 | 100 | NA | 2 | NA | 4 | 8 | NA |
| AZT+3TC+LPV/r | 62.5 | NA | NA | NA | NA | 8 | NA | NA | NA | NA |
| ABC+3TC+ATV/r | 57.1 | NA | NA | NA | NA | 7 | NA | NA | NA | NA |
| AZT+3TC+ATV/r | 60.0 | NA | NA | NA | NA | 5 | NA | NA | NA | NA |
| Other 2nd line | 50.0 | NA | NA | NA | NA | 2 | NA | NA | NA | NA |
| TDF+3TC+NVP | 100 | NA | NA | NA | NA | 2 | NA | NA | NA | NA |
| TDF+FTC+NVP | 100 | NA | NA | NA | NA | 2 | NA | NA | NA | NA |
| AZT+3TC+DTG | 100 | NA | NA | NA | NA | 1 | NA | NA | NA | NA |

Note: Values represent percentage suppressed and corresponding sample size for each regimen–year combination. Blank cells indicate regimen–year combinations not represented in the analytic cohort. Estimates for regimens with small sample sizes should be interpreted with caution.

**Supplementary Table S3: Transition Matrix with Distribution of Primary Regimen Optimization Pathways (N=30,414).**

This table details the most frequent transitions from baseline non-DTG regimens to optimized DTG-based therapy during the study period.

| Original Regimen (From) | Optimized Regimen (To) | Patient Count | Percentage (%) |
|---|---|---|---|
| TDF+3TC+EFV | TDF+3TC+DTG | 21,116 | **69.4%** |
| AZT+3TC+NVP | TDF+3TC+DTG | 3,164 | **10.4%** |
| AZT+3TC+EFV* | TDF+3TC+DTG | 1,638 | **5.4%** |
| TDF+FTC+EFV | TDF+3TC+DTG | 1,150 | **3.8%** |
| Others/Baseline Missing | TDF+3TC+DTG | 1,663 | **5.5%** |
| **Total Switchers** | | **30,414** | **100.0%** |

**Includes alternate coding for AZT+3TC+EFV and NVP-based backbones.*
